# Efficacy of Wang Nam Yen herbal tea on human milk production: A randomized controlled trial

**DOI:** 10.1101/2021.02.11.21251583

**Authors:** Koollachart Saejueng, Tanawin Nopsopon, Piyawadee Wuttikonsammakit, Wattanaporn Khumbun, Krit Pongpirul

## Abstract

**Background:** Insufficient milk production is a common problem affecting breastfeeding women, in particular following Cesarean delivery. Wang Nam Yen herbal tea is a promising traditional Thai medicine used by postpartum women to stimulate milk production, as an alternative to pharmaceutical galactagogues. We aimed to compare the efficacy of Wang Nam Yen herbal tea, domperidone, and placebo, in increasing milk production in mothers who underwent Cesarean delivery.

**Methods:** Women who underwent uncomplicated cesarean delivery at Sunpasitthiprasong Hospital were randomized into three groups. The participants received the treatments daily for three consecutive days. The primary outcome was breast milk volume at 72 hours after delivery. Secondary outcomes were pregnancy and neonatal outcomes, adverse events, and participant satisfaction.

**Results:** Of the 1,450 pregnant women that underwent cesarean delivery, 120 women were enrolled. Their mean age and gestational ages were 28.7 years and 38.4 weeks, respectively. Breast milk volume at 72 hours postpartum was significantly different among the three groups (*p* = 0.030). The posthoc Bonferroni correction indicated a significant difference in breast milk volume between Wang Nam Yen herbal tea group and placebo control group (*p* = 0.007) while there was no difference between Wang Nam Yen herbal tea group and domperidone group (*p* = 0.806) and between domperidone group and placebo control group (*p* = 0.018). There was no difference in pregnancy and neonatal outcomes, adverse events, and participant satisfaction among the three groups.

**Conclusion:** Wang Nam Yen herbal tea was effective in augmenting breast milk production at 72 hours postpartum in mothers following cesarean delivery, and there was no evidence that herbal tea and domperidone differed in terms of augmenting breast milk production.

**Trial registration:** The study was approved by the institutional review board of Sunpasithiprasong Hospital (No.061/2559) and was registered TCTR20170811003 with the Thai Clinical Trial Registry.

## Introduction

Breastfeeding advantages were well documented and considered the normative and recommended standard for infant nutrition [1, 2]. Low production of breast milk was one of the most common causes of inadequate breastfeeding, which could be supported through various techniques, including the use of substances that stimulate the production of breast milk, known as galactagogues [1, 2]. Domperidone was a commonly prescribed galactagogue, which increased breast milk production in women experiencing low breast milk supply, with low levels of transfer to maternal milk and no substantial alterations in the nutrient composition [3-6]. Despite no side-effects being reported in infants of mothers who had used domperidone, domperidone was controversial and had never been approved for marketing by the US FDA due to the potential increased risk of ventricular arrhythmia and sudden cardiac death in adults [2, 7, 8]. As an alternative to pharmaceutical products, several popular complementary and alternative techniques could be used as galactagogues [9]. Herbal medicines and techniques that had been found to support breastfeeding include herbal compresses to decrease engorgement pain, and herbal supplements, and commercially available herbal teas, such as those containing ginger, stinging nettle, fenugreek, and/or turmeric, and has shown to improve breast milk production without adverse effects [10-13]. In Thailand, numerous traditional galactagogues were commonly used, among those, banana flower, lemon basil, Thai basil, bottle gourd, and pumpkin had shown significant correlations with increased breast milk volume [14].

Several factors were associated with inadequate breastfeeding due to low milk production, including cesarean deliveries which can lead to lactation difficulties [1, 2, 15]. Postpartum treatment with Domperidone in women that have undergone full-term cesarean had shown augmented production of breast milk [15]. At Wang Nam Yen hospital in Thailand, the Wang Nam Yen herbal tea, consisting of sappan (*Caesalpinia sappan* Linn.), licorice (*Glycyrrhiza glabra* Linn.), bale fruit (*Aegle marmelos* L. Corr), ginger (*Zingiber officinale* Roscoe), and jewel vine (*Glycyrrhiza glabra* Linn.) had been used as a traditional Thai medicine to stimulate milk production in postpartum women. Acute oral toxicity assessment of the tea in Wistar rats revealed a median lethal dose (LD50) of 5,000 mg/kg with no cases of serious side effects. While observational data from the hospital found correlations in breast milk volume with the use of the herbal tea, with no serious side effects, a randomized controlled trial was necessary to provide substantive evidence of the herbal tea efficacy in stimulating breast milk production.

The Wang Nam Yen herbal tea was a promising traditional Thai galactagogue, which might be used as an alternative to the common pharmaceutical galactagogue, domperidone, to support breastfeeding difficulties due to low supply, especially in women that have undergone cesarean deliveries. The objective of this study was to compare the efficacy of Wang Nam Yen herbal tea, domperidone, and a placebo, in augmenting breast milk production in mothers that have undergone Cesarean delivery, in a double-blinded randomized controlled trial.

## Methods

### Study design

Tea4Milk was a three-armed, double-blinded randomized controlled trial on human milk production conducted in Ubon Ratchathani Province, Thailand at Sunpasithiprasong Hospital, a regional hospital under the Ministry of Public Health, from February 2017 to September 2017. Participants were recruited by placing advertisements in four obstetrics wards. All patients provided written informed consent. The study was reviewed and approved by the institutional review board of Sunpasithiprasong Hospital (No.061/2559). The trial was registered and kept up to date at the Thai Clinical Trial Registry (Registration No. TCTR20170811003).

### Participants

Inclusion criteria were women aged 15 to 41 years that had delivered via cesarean section at 28 to 42 weeks’ gestational age. Participants with contraindications for breastfeeding and/or serious illness affecting mother or infant, such as human immunodeficiency virus infection, postpartum hemorrhage with hypovolemic shock, HELLP syndrome (hemolysis, elevated liver enzymes, and low platelet count), eclampsia with respiratory failure, or had a history of allergy to domperidone or ingredients in the herbal tea were excluded from the study.

### Randomization and blinding

Participants were randomly assigned into three groups based on numbers generated by a table of random numbers in blocks of three: 1) Wang Nam Yen herbal tea and placebo tablet (T group), 2) domperidone tablet and placebo herbal tea (D group), 3) placebo tablet and placebo herbal tea (C group). All study interventions were prepacked and sealed in opaque packages, for three consecutive days of postpartum use. Participants and investigators were blinded to the treatment assignment.

### Interventions

Wang Nam Yen herbal tea was manufactured by Wang Nam Yen hospital, a community hospital in Sa Kaeo reputed for Thai traditional herbal medicine. Each Wang Nam Yen herbal tea contained sappan (*Caesalpinia sappan* Linn.), licorice (*Glycyrrhiza glabra* Linn.), bale fruit (*Aegle marmelos* L. Corr), ginger (*Zingiber officinale* Roscoe), and jewel vine (*Glycyrrhiza glabra* Linn.). Wang Nam Yen herbal tea was packed in a tea bag designed for each meal. The herbal tea placebo was also prepared as a teabag for each meal, and the placebo tea bag was identical to Wang Nam Yen teabag. Both herbal teas were prepared by diffusion in 200 mL of warm water for 5 minutes. Interventions were administered orally three times per day after meals. The first administration occurred 12-18 hours following delivery to ensure sufficient time for the evaluation of any postpartum complications that may have occurred. Domperidone was administered at 10 mg per dose, as 30 mg of domperidone per day was considered safe and sufficient for increasing breast milk production [16, 17]. The placebo tablet was identical to the appearance of the domperidone tablet. All participants (T, D, C groups) were administered 200 mL of tea and a tablet after each meal.

T–The Wang Nam Yen herbal tea treatment group received Wang Nam Yen herbal tea and placebo tablet after each meal for three meals per day. This was an intervention arm that was designed to assess the efficacy of Wang Nam Yen herbal tea.

D–The domperidone treatment group received an herbal tea placebo and a 10 mg domperidone tablet after each meal for three meals per day. This was an active control arm that was designed to be a comparator as standard treatment.

C–The placebo control group received both herbal tea placebo and a placebo tablet after each meal for three meals per day. This was a placebo control arm that was designed to be a reference group with no treatment.

### Outcome measures

The primary outcome was breast milk volume at 72 hours after delivery. All participants were encouraged to breastfeed their infant within 24 hours of delivery based on Sanpasitthiprasong hospital’s standard of care policy of early breastfeeding, frequent breastfeeding, correct positioning, and avoiding the use of formula when possible. The breast milk volume was measured with a two-hour interval from the last breastfeeding, using an electronic pump (Spectra S2Plus^®^, Spectra Baby USA). The electronic pump was applied for 15 minutes on each breast and the total volume of milk was recorded. Breast milk volume was also measured at 24 hours and 48 hours post-delivery. The breast milk volume at 24 hours post-delivery was used as the baseline.

Secondary outcomes were observed 72 hours after the delivery period including (1) pregnancy outcomes such as postpartum hemorrhage, endometritis, (2) neonatal outcomes such as neonatal jaundice, respiratory distress, (3) adverse effects including headache, dry mouth, diarrhea, muscle cramps, itching, or allergic reactions, and (4) participant satisfaction during the three days’ postpartum period were also recorded.

### Sample size calculation

We used mean and standard deviation of breast milk volume at day 4 after delivery based on the study by Jantarasaengaram and Sreewapa, 2012 on the effects of domperidone compared to placebo in augmenting lactation following cesarean delivery with 80% power and a 2-sided type I error at 5% [15]. With taking into account an expected follow-up loss of 15%, the anticipated sample size of 120 would require a sample size of 40 in each group.

### Statistical analysis

The statistical analysis was performed on an intention-to-treat basis using Stata/MP software version 15.0 (StataCorp 2017, College Station, TX). Descriptive statistics were carried out using mean with standard deviation or median with interquartile range when appropriate. Categorical data were presented as counts and percentage and tested for significance with the Chi-square test. Continuous data were assessed for normality and tested for significance with one-way ANOVA with Bonferroni correction and Kruskal-Wallis tests for comparison among groups. Statistical significance was defined as *p* < 0.05.

## Results

### Participant characteristics

The 1450 women who underwent Cesarean delivery in Sunpasitthiprasong hospital from February 2017 to September 2017 were assessed for eligibility. 303 were excluded due to severe eclampsia, eclampsia, HELLP syndrome, or neonatal separation from mother immediately after birth and 1027 declined participation. A total of 120 mother-infant dyads were enrolled in this study and randomized into three groups, with 40 participants in each (Fig 1). There were six participants whose infants required special care in the neonatal intensive care unit but continued to express their milk via breast pump; three were from the control group, two from the domperidone group, and one from the herbal tea group.

**Fig 1.**
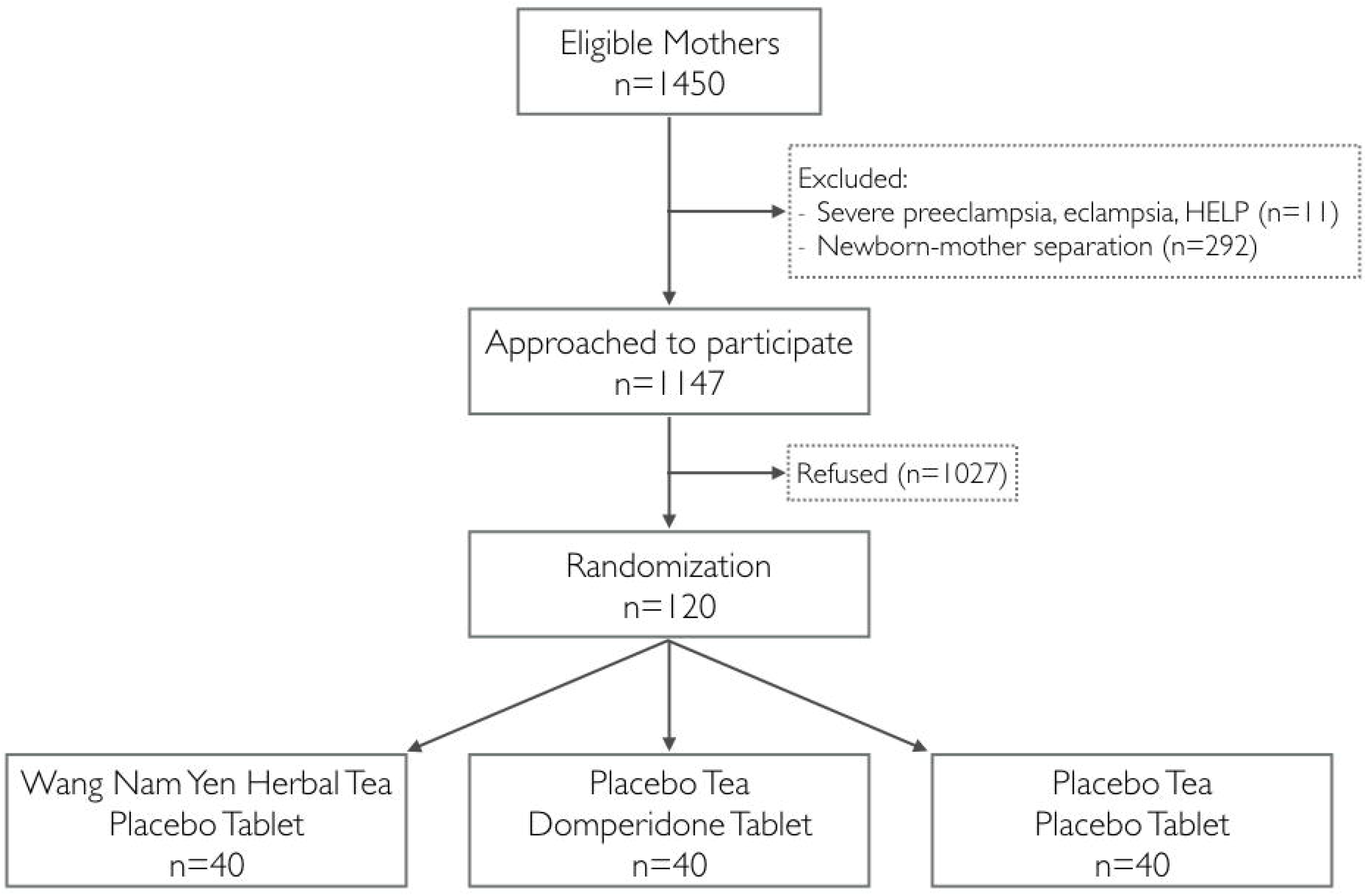
Flow diagram of the clinical trial.

The average participant age was 28.7 years old with an average gestational age at delivery of 38.4 weeks. There were no significant differences among the groups in demographic data including maternal age, occupation, highest education, method of payment, and salary per month; or in baseline obstetric data including gestational age, parity, the number of antenatal visits, labor symptoms, the indication of cesarean delivery, birth weight, sex of the baby and immediate postpartum hemorrhage defined by blood loss more than 1000ml. There were no significant differences between the groups in the average fluid intake and output during the 72 hours postpartum period or the amount of formula milk required (Table 1).

**Table 1.** Baseline characteristics of the study participants.

| Baseline data | Total (n=120) | Herbal tea (n=40) | Domperidone (n=40) | Control (n=40) | p value |
| --- | --- | --- | --- | --- | --- |
| Demographic characteristics |  |  |  |  |  |
| Age (years; mean ± SD) | 28.7 ± 5.9 | 29.2 ± 5.7 | 28.7 ± 5.9 | 28.3 ± 6.2 | 0.804 |
| Occupation, n (%) |  |  |  |  | 0.160 |
| Self-employed | 19 (15.8%) | 4 (10.0%) | 4 (10.0%) | 11 (27.5%) |  |
| Farmer | 10 (8.3%) | 2 (5.0%) | 5 (12.5%) | 3 (7.5%) |  |
| Employee | 41 (34.2%) | 16 (40.0%) | 13 (32.5%) | 12 (30.0%) |  |
| Government officials | 8 (6.7%) | 5 (12.5%) | 3 (7.5%) | 0 (0.0%) |  |
| Housewife | 41 (34.2%) | 12 (30.0%) | 15 (37.5%) | 14 (35.0%) |  |
| Others | 1 (0.8%) | 1 (2.5%) | 0 (0.0%) | 0 (0.0%) |  |
| Monthly income (baht; median, IQR) (n=99)* | 5000 (0, 14000) | 5000 (0, 15000) | 3000 (0, 11500) | 3600 (0, 13000) | 0.682 |
| Method of payment, n (%) |  |  |  |  | 0.283 |
| Public universal health coverage | 67 (55.8%) | 20 (50.0%) | 27 (67.5%) | 20 (50.0%) |  |
| Social welfare | 37 (30.8%) | 13 (32.5%) | 11 (27.5%) | 13 (32.5%) |  |
| Government official's welfare | 8 (6.7%) | 4 (10.0%) | 2 (5.0%) | 2 (5.0%) |  |
| Other | 8 (6.7%) | 3 (7.5%) | 0 (0.0%) | 5 (12.5%) |  |
| Highest education, n (%) |  |  |  |  | 0.419 |
| Elementary school | 13 (10.8%) | 5 (12.5%) | 6 (15.0%) | 2 (5.0%) |  |
| Secondary school | 63 (52.5%) | 22 (55.0%) | 19 (47.5%) | 22 (55.0%) |  |
| Vocational diploma | 12 (10.0%) | 1 (2.5%) | 5 (12.5%) | 6 (15.0%) |  |
| Bachelor's degree or higher | 32 (26.7%) | 12 (30.0%) | 10 (25.0%) | 10 (25.0%) |  |
| Obstetric characteristics |  |  |  |  |  |
| Gestational age (weeks; mean ± SD) | 38.4 ± 1.5 | 38.6 ± 1.0 | 38.1 ± 2.0 | 38.7 ± 1.1 | 0.142 |
| Primigravida, n (%) | 41 (34.2%) | 12 (30.0%) | 12 (30.0%) | 17 (42.5%) | 0.396 |
| Antenatal visits more than 3 times, n (%) | 58 (48.3%) | 19 (47.5%) | 19 (47.5%) | 20 (50.0%) | 0.967 |
| Presence of labor symptoms, n (%) | 66 (55.0%) | 21 (52.5%) | 23 (57.5%) | 22 (55.0%) | 0.904 |
| Indication of Cesarean delivery, n (%) |  |  |  |  | 0.946 |
| Cephalopelvic disproportion | 48 (40.0%) | 15 (37.5%) | 15 (37.5%) | 18 (45.0%) |  |
| Fetal distress | 8 (6.7%) | 3 (7.5%) | 3 (7.5%) | 2 (5.0%) |  |
| Abnormal presentation | 9 (7.5%) | 3 (7.5%) | 4 (10.0%) | 2 (5.0%) |  |
| Previous Cesarean delivery | 48 (40.0%) | 18 (45.0%) | 15 (37.5%) | 15 (37.5%) |  |
| Others | 7 (5.8%) | 1 (2.5%) | 3 (7.5%) | 3 (7.5%) |  |
| Experience of breastfeeding, n (%) | 64 (53.3%) | 24 (60.0%) | 21 (52.5%) | 19 (47.5%) | 0.529 |
| Nurturing environment, n (%) |  |  |  |  | 0.699 |
| Single mom | 1 (0.8%) | 0 (0.0%) | 1 (2.5%) | 0 (0.0%) |  |
| Single family | 47 (39.2%) | 15 (37.5%) | 15 (37.5%) | 17 (42.5%) |  |
| Extended family | 72 (60.0%) | 25 (62.5%) | 24 (60.0%) | 23 (57.5%) |  |
| Birthweight (grams; median, IQR) | 3067.5<br>(2737.5, 3337.5) | 3147.5<br>(2736.3, 3475.0) | 3055.0<br>(2743.8, 3302.5) | 2975.0<br>(2715.0, 3307.5) | 0.396 |
| Sex of baby, n (%) |  |  |  |  | 0.670 |
| Female | 60 (50.0%) | 20 (50.0%) | 18 (45.0%) | 22 (55.0%) |  |
| Male | 60 (50.0%) | 20 (50.0%) | 22 (55.0%) | 18 (45.0%) |  |
| Immediate PPH, n (%) | 2 (1.7%) | 1 (0.8%) | 0 (0.0%) | 1 (0.8%) | 0.601 |
| Fluid status during 72 hours postpartum period |  |  |  |  |  |
| Average fluid intake per day (ml) | 2566.2 ± 560.6 | 2525.7 ± 436.9 | 2602.9 ± 573.6 | 2569.9 ± 660.4 | 0.828 |
| Average fluid output per day (ml) | 1826.0 ± 542.7 | 1781.3 ± 452.5 | 1812.6 ± 583.2 | 1884.2 ± 590.0 | 0.689 |

Statistics were analyzed by using one-way ANOVA for normally distributed continuous variables, Kruskal-Wallis test for non-normal distributed continuous variables, and Chi-square test for categorical variables. IQR, interquartile range; PPH, postpartum hemorrhage; SD, Standard deviation
\*Data not available for all randomized participants

### Primary and secondary outcomes

Breast milk volume at 72 hours postpartum was significantly different among the three groups (*p* = 0.030). The posthoc Bonferroni correction indicated a significant difference in breast milk volume between Wang Nam Yen herbal tea group and placebo control group (*p* = 0.007) while there was no difference between Wang Nam Yen herbal tea group and domperidone group (*p* = 0.806), and between domperidone group and placebo control group (*p* = 0.018) (Fig 2). While there was a significant difference in baseline breast milk volume at 24 hours postpartum among the three groups (*p* = 0.036), the significant difference between groups was not observed with Bonferroni correction: *p* = 0.039 for herbal tea and control groups, *p* = 0.433 for domperidone and control groups, and *p* = 0.089 for herbal tea and domperidone groups. There was no significant breast milk volume observed among the three groups at 48 hours postpartum (*p* = 0.256).

**Fig 2.**
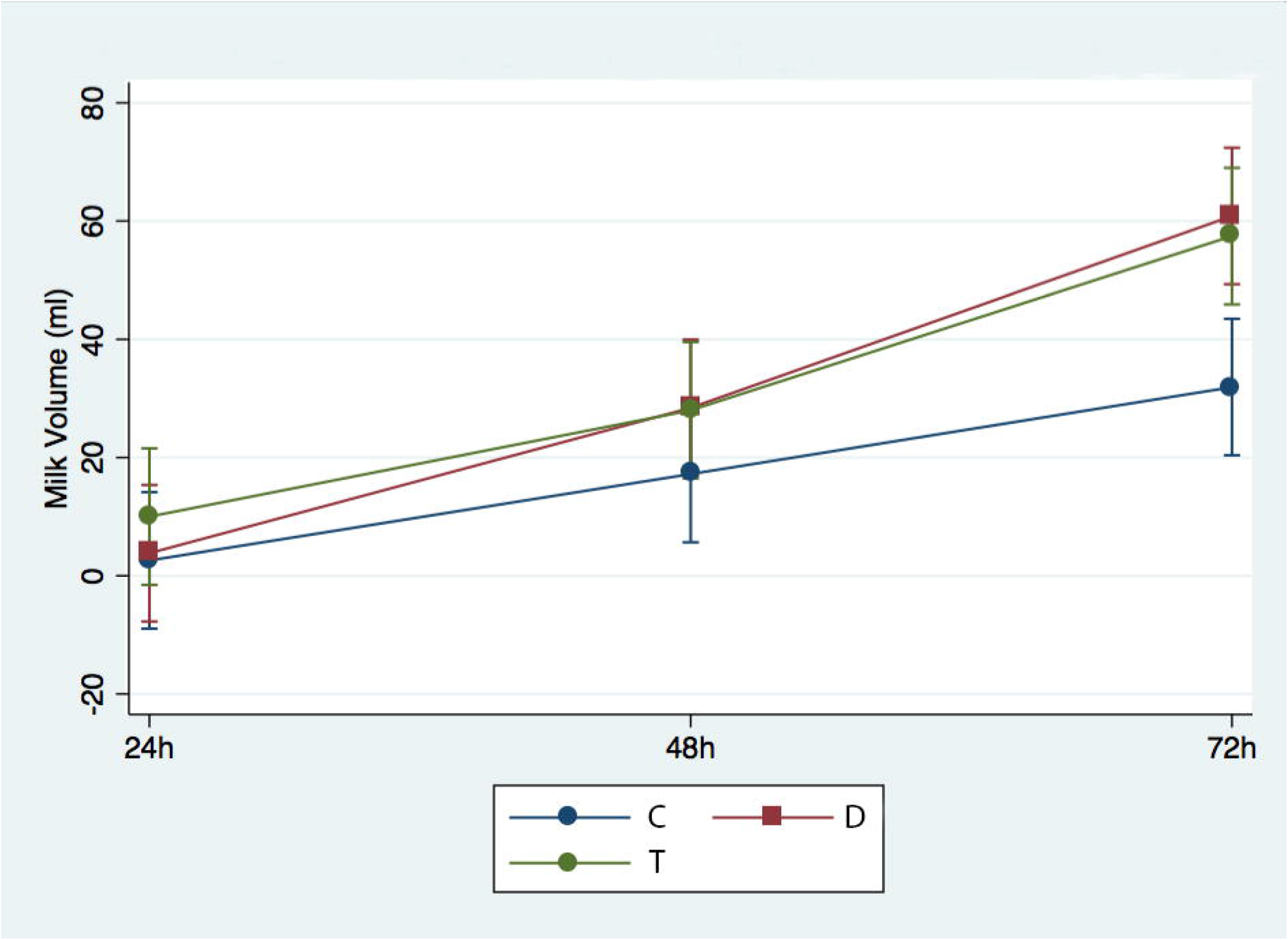
Change in human milk volume over time. Mean and standard deviation are presented. C, control group; D, domperidone group; T, herbal tea group.

There were no significant differences in maternal or neonate complications among the three groups (Table 2). One participant in the control group developed postpartum endometritis, one participant in the domperidone group reported dry mouth and one participant in the herbal tea group reported diarrhea as an adverse effect during the 72 hours. Six infants developed jaundice, two in the control and four in the herbal tea group, and six required neonatal intensive care unit admission, three in the control, two in the domperidone, and one in the herbal tea group.

**Table 2.** Secondary outcome measures for maternal complications, neonatal complications, and drug adverse events.

| Secondary outcomes | Total (n=120) | Herbal tea (n=40) | Domperidone (n=40) | Control (n=40) | p value |
| --- | --- | --- | --- | --- | --- |
| Maternal complications |  |  |  |  |  |
| Late PPH | 0 (0.0%) | 0 (0.0%) | 0 (0.0%) | 0 (0.0%) | N/A |
| Postpartum endometritis | 1 (0.8%) | 0 (0.0%) | 0 (0.0%) | 1 (2.5%) | 0.365 |
| Neonatal complications |  |  |  |  |  |
| Neonatal jaundice | 6 (5.0%) | 4 (10.0%) | 0 (0.0%) | 2 (5.0%) | 0.127 |
| NICU admission | 6 (5.0%) | 1 (2.5%) | 2 (5.0%) | 3 (7.5%) | 0.591 |
| Drug adverse events |  |  |  |  |  |
| Adverse event at 24 hours | 2 (1.7%) | 1 (2.5%) | 1 (2.5%) | 0 (0.0%) | 0.601 |
| Adverse event at 48 hours | 2 (1.7%) | 1 (2.5%) | 1 (2.5%) | 0 (0.0%) | 0.601 |
| Adverse event at 72 hours | 2 (1.7%) | 1 (2.5%) | 1 (2.5%) | 0 (0.0%) | 0.601 |

Statistics were analyzed by the Chi-square test. Drug adverse event includes headache, dry mouth, diarrhea, muscle cramps, itching, or allergic reactions. NICU, neonatal intensive care unit; N/A, not applicable; PPH, postpartum hemorrhage.

Based on participant satisfaction outcomes, 64.2% of all the participants liked their breastfeeding experience during the study period, 71.7% liked using galactagogues to increase their milk production and 75.8% would use galactagogues in their subsequent pregnancy, while 18.3% of participants worried about the adverse effects of using galactagogues (Table 3). While not significantly different between the intervention groups, more participants in the control group (32.5%) reported having problems with breastfeeding than participants in the domperidone (20.0%) and herbal tea group (17.5%), and also more participants in herbal tea (62.5%) and domperidone (55.0%) groups agreed to continue the use of galactagogue post-study compare to the control group (35.0%). 68.3% of participants would recommend galactagogues to a friend, 75% would recommend food to enhance milk production and 78.3% would recommend behavior modifications to enhance milk production.

**Table 3.** Secondary outcome measures for participant satisfaction.

| Questions | Total<br>(n= 120) | Herbal tea<br>(n=40) | Domperidone<br>(n=40) | Control<br>(n=40) | *p* value |
| --- | --- | --- | --- | --- | --- |
| 1. You like your breastfeeding |  |  |  |  | 0.835 |
| Agree | 77 (64.2%) | 28 (70.0%) | 24 (60.0%) | 25 (62.5%) |  |
| Disagree | 4 (3.3%) | 1 (2.5%) | 1 (2.5%) | 2 (5.0%) |  |
| Not sure | 39 (32.5%) | 11 (27.5%) | 15 (37.5%) | 13 (32.5%) |  |
| 2. You like to use this galactagogue |  |  |  |  | 0.143 |
| Agree | 86 (71.7%) | 29 (72.5%) | 28 (70.0%) | 29 (72.5%) |  |
| Disagree | 3 (2.5%) | 0 (0.0%) | 0 (0.0%) | 3 (7.5%) |  |
| Not sure | 31 (25.8%) | 11 (27.5%) | 12 (13.0%) | 8 (20.0%) |  |
| 3. You have a problem with your breastfeeding |  |  |  |  | 0.200 |
| Agree | 28 (23.3%) | 7 (17.5%) | 8 (20.0%) | 13 (32.5%) |  |
| Disagree | 37 (30.8%) | 17 (42.5%) | 10 (25.0%) | 10 (25.0%) |  |
| Not sure | 55 (45.8%) | 16 (40.0%) | 22 (55.0%) | 17 (42.5%) |  |
| 4. You worried about galactagogues adverse effect |  |  |  |  | 0.662 |
| Agree | 22 (18.3%) | 5 (12.5%) | 7 (17.5%) | 10 (25.0%) |  |
| Disagree | 56 (46.7%) | 19 (47.5%) | 20 (50.0%) | 17 (42.5%) |  |
| Not sure | 42 (35.0%) | 16 (40.0%) | 13 (32.5%) | 13 (32.5%) |  |
| 5. You decided to continue using galactagogue |  |  |  |  | 0.132 |
| Agree | 61 (50.8%) | 25 (62.5%) | 22 (55.0%) | 14 (35.0%) |  |
| Disagree | 10 (8.3%) | 3 (7.5%) | 2 (5.0%) | 5 (12.5%) |  |
| Not sure | 49 (40.8%) | 12 (30.0%) | 16 (40.0%) | 21 (52.5%) |  |
| 6. You will recommend your friend to use this galactagogue |  |  |  |  | 0.171 |
| Agree | 82 (68.3%) | 29 (72.5%) | 27 (67.5%) | 26 (65.0%) |  |
| Disagree | 3 (2.5%) | 0 (0.0%) | 0 (0.0%) | 3 (7.5%) |  |
| Not sure | 35 (29.2%) | 11 (27.5%) | 13 (32.5%) | 11 (27.5%) |  |
| 7. You will recommend your friend to eat food enhancing breast milk production |  |  |  |  | 0.791 |
| Agree | 90 (75.0%) | 31 (77.5%) | 28 (70.0%) | 31 (77.5%) |  |
| Disagree | 4 (3.3%) | 1 (2.5%) | 1 (2.5%) | 2 (5.0%) |  |
| Not sure | 26 (21.7%) | 8 (20.0%) | 11 (27.5%) | 7 (17.5%) |  |
| 8. You will advise your friend on behavioral ways to continue breastfeeding |  |  |  |  | 0.942 |
| Agree | 94 (78.3%) | 32 (80.0%) | 30 (75.0%) | 32 (80.0%) |  |
| Disagree | 5 (4.2%) | 2 (5.0%) | 2 (5.0%) | 1 (2.5%) |  |
| Not sure | 21 (17.5%) | 6 (15.0%) | 8 (20.0%) | 7 (17.5%) |  |
| 9. You will use galactagogue in your subsequent pregnancy |  |  |  |  | 0.412 |
| Agree | 91 (75.8%) | 32 (80.0%) | 30 (75.0%) | 29 (72.5%) |  |
| Disagree | 5 (4.2%) | 2 (5.0%) | 0 (0.0%) | 3 (7.5%) |  |
| Not sure | 24 (20.0%) | 6 (15.0%) | 10 (25.0%) | 8 (20.0%) |  |

Statistics were analyzed by the Chi-square test.

## Discussion

In this study, Wang Nam Yen herbal tea effectively increased breast milk volume production at 72 hours postpartum compared to placebo. There was no significant difference in breast milk volume between Wang Nam Yen herbal tea and the domperidone group. The difference in early augmentation effect at 24 hours postpartum was observed among three groups, but there was no significant difference between any two groups when applying Bonferroni correction. Wang Nam Yen herbal tea and domperidone had no serious side effects on the mothers or infants. One participant reported diarrhea and one participant reported dry mouth as adverse effects from the herbal tea and domperidone group, respectively. The overall satisfaction of using galactagogues was high and most participants would recommend using galactagogues to friends for increasing milk volume production.

Although the study did not measure prolactin levels, the findings demonstrate the efficacy of the herbal tea in enhancing milk volume. While the method of collecting milk, using a breast pump to measure milk volume, might affect the study outcome as electronic pumps can disturb normal biological synthesis of breast milk, the results showed the same trend as previous studies of domperidone at 30 mg/day dose in the efficacy to increase breast milk volume [3, 5, 15, 16, 18]. However, breast milk volume at 72 hours postpartum in the domperidone group showed no significant difference from the placebo control group when using Bonferroni correction. This might occur from the fact that Bonferroni’s correction was too conservative, thus reduce the power of the study. The hospital’s standard policy for breastfeeding, including encouraging early breastfeeding, frequent breastfeeding, correct position, and avoiding the use of the formula is important recommendations that should be strictly followed by all mothers before using food, behavior modifications, or galactagogues to increase milk volume production. The findings support the use of galactagogues in addition to behavioral recommendations and food intake to increase breast milk production in mothers with insufficient milk volume following a cesarean delivery [14].

In terms of safety, the side effects of domperidone could include dry mouth, headache, insomnia, abdominal cramps, diarrhea, nausea, and urinary retention [15]. The study found no major side effects, with one report of dry mouth as an adverse effect of domperidone and one report of diarrhea as an adverse effect of the herbal tea, which subsided with discontinued use of the intervention.

Supporting findings from previous studies, there were no significant differences in neonatal outcomes, especially neonatal jaundice, between the intervention and control groups, however, more research may be needed to conclusively support these results [2]. There were no maternal complications reported, including postpartum endometritis and postpartum hemorrhage, as a result of using domperidone or herbal tea. Based on the study, there are no safety concerns in using herbal tea during the puerperal period.

As a randomized controlled trial, one of the strengths of this study was by design, as careful randomization and blinding minimized potential confounding factors and biases. This trial was among the first studies to examine the effects of Wang Nam Yen tea compared to domperidone and a placebo. Although some participants declined to enroll in the trial following randomization, the sample size of this study provided sufficient power and was able to determine the efficacy of Wan Nam Yen herbal tea in increasing breast milk production at 72 hours postpartum as the primary outcome. Therefore, the results of this study could be applied to pregnant women that underwent cesarean delivery at 28 to 40-week gestational age.

A limitation of this study is that the efficacy of the interventions on other outcomes, such as prolactin level and breast milk volume at 7 days post-administration, to confirm hormonal effect and maintenance of augmentation, was not evaluated.

## Conclusion

Wang Nam Yen herbal tea, an alternative traditional Thai medicine, was effective in augmenting breast milk production at 72 hours postpartum in mothers following cesarean delivery, and there was no evidence that herbal tea and domperidone differed in terms of augmenting breast milk production.

## Supporting information

CONSORT Checklist

## Data Availability

All data relevant to the study are included in the article. De-identified data of this study could be shared upon request.

## Acknowledgments

The authors thank Mr. Pinit Chinsoi, Wang Nam Yen hospital, for preparing the herbal and placebo teas, and the Faculty of Pharmaceutical Sciences, Ubon Ratchathani University for preparing the Domperidone. We thank Dr. Charnsak Jungmunkong and contributors from Sunpasitthiprasong Hospital for the administrative support of this trial.

## Author Contributions

**Conceptualization:** Koollachart Saejueng, Krit Pongpirul

**Data curation:** Koollachart Saejueng, Piyawadee Wuttikonsammakit, Krit Pongpirul

**Formal analysis:** Koollachart Saejueng, Tanawin Nopsopon

**Funding acquisition:** Wattanaporn Khumbun, Krit Pongpirul

**Investigation:** Koollachart Saejueng, Tanawin Nopsopon, Piyawadee Wuttikonsammakit, Wattanaporn Khumbun, Krit Pongpirul

**Methodology:** Koollachart Saejueng, Tanawin Nopsopon, Krit Pongpirul

**Project administration:** Koollachart Saejueng, Wattanaporn Khumbun

**Supervision:** Krit Pongpirul

**Writing – original draft:** Koollachart Saejueng, Tanawin Nopsopon, Krit Pongpirul

**Writing – review & editing:** Koollachart Saejueng, Tanawin Nopsopon, Piyawadee Wuttikonsammakit, Wattanaporn Khumbun, Krit Pongpirul

## Funding

The authors received financial support for the research from the Department of Thai Traditional and Alternative Medicine, Ministry of Public Health, Thailand. The funding source had no involvement in research preparation, study design; collection, analysis, interpretation of data; writing of the report; or in the decision to submit the article for publication.

## Competing interests

The authors have declared that no competing interests exist.

## Supporting information

**S1 File. CONSORT checklist**.

(PDF)

## Notes

### Competing Interest Statement

The authors have declared no competing interest.

### Clinical Trial

TCTR20170811003

### Author Declarations

The study was approved by the institutional review board of Sunpasithiprasong Hospital (No.061/2559).

